# Isolate differences in colonization efficiency during experimental human pneumococcal challenge

**DOI:** 10.1101/2020.04.20.20066399

**Authors:** Sherin Pojar, Alan Basset, Jenna F. Gritzfeld, Elissavet Nikolaou, Saskia van Selm, Marc J. Eleveld, Rebecca A. Gladstone, Carla Solórzano, Ankur B. Dalia, Esther German, Elena Mitsi, Victoria Connor, Angela D. Hyder-Wright, Helen Hill, Caz Hales, Tao Chen, Andrew Camilli, Andrea M. Collins, Jamie Rylance, Stephen D. Bentley, Simon P. Jochems, Marien I. de Jonge, Jeffrey N. Weiser, David W. Cleary, Stuart Clarke, Richard Malley, Stephen B. Gordon, Daniela M. Ferreira

## Abstract

Colonization efficiency varies considerably between *Streptococcus pneumoniae* (pneumococcus) strains. The microbial characteristics that influence those differences are still largely unknown. Here, we report rates and kinetics of colonization of four pneumococcal strains upon experimental human pneumococcal challenge. Healthy adults were intranasally challenged with one of four pneumococcal strains (serotype/clonal name: 6B/BHN418, 15B/SH8286, 23F/P1121 and 23F/P833) over a range of doses. Maximum colonization achieved was 60%, 31%, 16% and 10%, respectively. Density and duration of colonization did not differ significantly between the tested strains. We further evaluated murine colonization, non-opsonic neutrophil mediated killing, epithelial cell adherence and average chain length of these four pneumococcal strains. Of these, only chain length was found to be associated with colonization efficiency in the human challenge model. Our data demonstrate that colonization rates following experimental challenge vary with the strain used and suggest that efficiency in colonization is related to pneumococcal chain length.

## Introduction

Colonization of the nasopharynx is the first and obligatory step in pneumococcal pathogenesis and the reservoir for transmission in the population (Simell et al., 2012). Colonization rates vary considerably between pneumococcal serotypes as well as between community groups. Host factors known to affect susceptibility to colonization include age, geographic area, socio-economic status, and innate and adaptive immune responses (Goldblatt et al., 2005, Lu et al., 2008, Bogaert et al., 2009, Principi et al., 1999, Garcia-Rodriguez and Fresnadillo Martinez, 2002, Glennie et al., 2016, Pennington et al., 2016). Microbial factors influencing pneumococcal colonization patterns are less well understood. Pneumococcal capsule plays a large role in determining colonization as well as transmission efficiency (Zafar et al., 2017, Trzcinski et al., 2015, Nelson et al., 2007, Lees et al., 2017). Further, factors potentially influencing successful colonization are microbial properties contributing to epithelial adherence, phase variation and interaction with the resident microflora (Simell et al., 2012). Identifying factors that influence pneumococcal patterns of colonization is important to public health and may ultimately allow the development of targeted intervention methods as well as better understanding of epidemiological surveillance data.

Experimental human pneumococcal challenge (EHPC) is a well-established and controlled model for studying host immunity and host-pathogen interactions as well as pneumococcal colonization dynamics, since the precise bacterial dose, timing of colonization onset and duration are known (Jochems et al., 2018, Glennie et al., 2016, Pennington et al., 2016, Mitsi et al., 2017, Wright et al., 2013, Ferreira et al., 2013). Here, we compare the rates and kinetics of colonization of four different pneumococcal strains using the EHPC model. These strains are further evaluated in murine models to determine if the animal model can be used to predict pneumococcal colonization efficiency in humans. Moreover, we test key microbiological properties which vary by capsular type and have been associated with pneumococcal colonization prevalence: non-opsonic neutrophil mediated killing (Weinberger et al., 2009), adherence to human nasal epithelial cells (Keller et al., 2013) and pneumococcal chain length (Rodriguez et al., 2012).

## Results

### EHPC study characteristics

We recruited and intranasally challenged a total of 190 volunteers in four dose-ranging studies. Sixty volunteers were challenged with serotype 6B clone BHN418 (6B/BHN418), 60 with serotype 23F clone P833 (23F/P833), 16 with serotype 23F clone P1121 (23F/P1121) and 54 with serotype 15B clone SH8286 (15B/SH8286). Doses administered ranged from 10,000 CFU/nostril to 320,000 CFU/nostril. Age and gender distribution were comparable between the studies (Table 1). There were no reported severe adverse events secondary to pneumococcal challenge in these studies.

**Table 1.** Demographics.

| **Pneumococcal strain** | **Median Age -yr (range)** | **Female - no. (%)** |
| --- | --- | --- |
| **6B/BHN418** | 22 (18-58) | 32/60 (53%) |
| **15B/SH8286** | 21 (18-49) | 31/54 (57%) |
| **23F/P1121** | 29 (25-47) | 11/16 (69%) |
| **23F/P833** | 21 (18-45) | 33/60 (55%) |

### Dose-response relationship between pneumococcal strains upon human challenge

The dose-response relationship upon human challenge differed between the tested strains as shown in Figure 1. Notably, strains 23F/P833, 23F/P1121, and 15B/SH8286 showed declining colonization rates for doses higher than 8×10^4^ CFU/nostril. This was not the case for strain 6B/BHN418 which achieved maximum colonization rates at doses higher than 8×10^4^ CFU/nostril.

**Figure 1:**
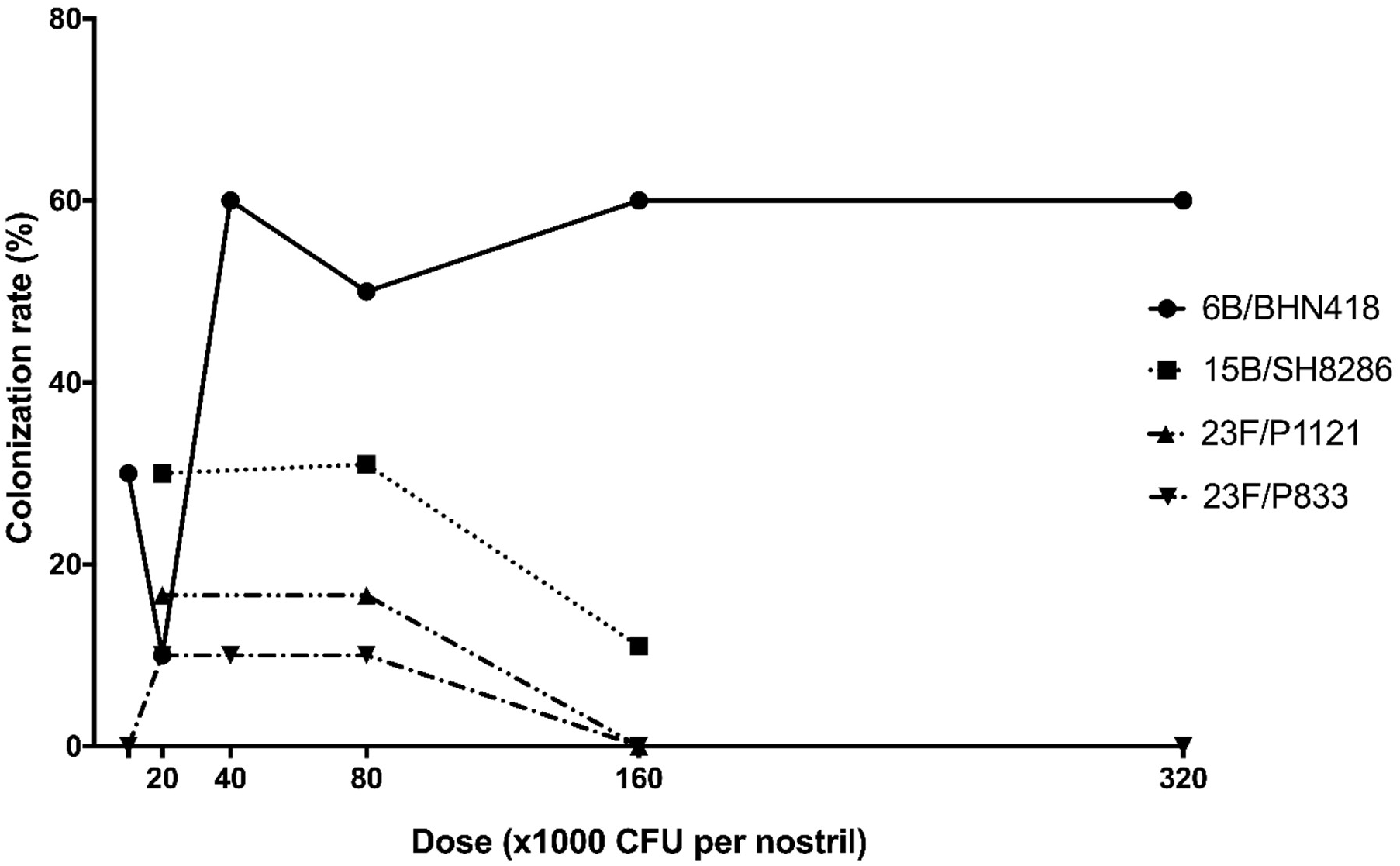
Dose-response relationship between pneumococcal strains upon human challenge. Volunteers were intranasally challenged with either 6B/BHN418 (circle), 15B/SH8286 (square), 23F/P1121 (triangle apex up) or 23F/P833 (triangle apex down). Each point represents the percentage of colonization acquisition achieved with the specified pneumococcal strain when volunteers were exposed with 1 × 10^4^, 2 × 10^4^, 4 × 10^4^, 8 × 10^4^, 16 × 10^4^ and 32 × 10^4^ pneumococci per nostril. Colonization was determined by growth of the specified pneumococcal strain on Columbia blood agar from nasal wash at any time point following challenge.

Maximum colonization achieved was 60% (6/10) for 6B/BHN418 with 4×10^4^, 16×10^4^ and 32×10^4^ CFU/nostril, 31% (11/35) for 15B/SH8286 with 8×10^4^ CFU/nostril, 16% (1/6) for 23F/P1121 with 2×10^4^ and 8×10^4^ CFU/nostril, while 23F/P833 achieved maximum colonization of 10% (1/10) with 2×10^4^, 4×10^4^ and 8×10^4^ CFU/nostril.

### Pneumococcus 6B clone BHN418 shows highest colonization rate upon human challenge

Colonization rates averaged over all doses (Table 2) differed significantly between the tested strains (Chi-square test, P<0.0001). Most efficient colonization was observed with 6B/BHN418 (27/60, 45%). A trend towards less efficient colonization was observed with 15B/SH8286 (15/54, 28%) when compared to 6B/BHN418 (P=0.0570). Colonization efficiency of 23F/P1121 (2/16, 13%) and 23F/P833 (3/60, 5%) were comparable (P=0.2823) but were significantly lower than that of 6B/BHN418 (P=0.0174 and P<0.0001, respectively).

**Table 2.**
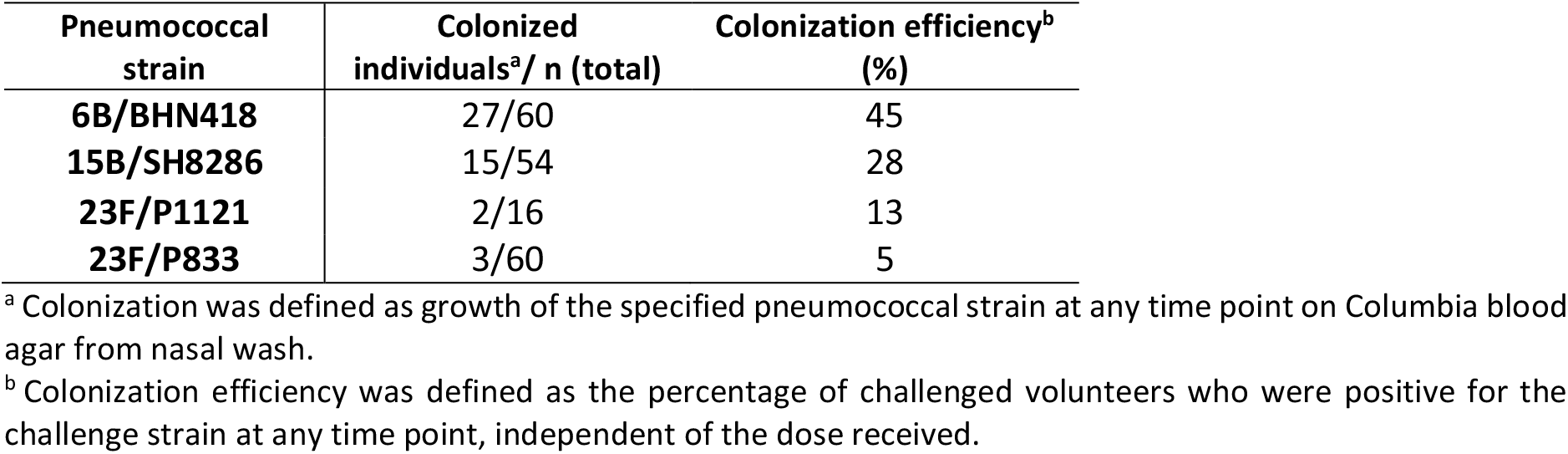
Pneumococcus 6B clone BHN418 shows highest colonization rates upon human challeng*e*.

| **Pneumococcal strain** | **Colonized individuals<sup>a</sup>/ n (total)** | **Colonization efficiency<sup>b</sup> (%)** |
| --- | --- | --- |
| **6B/BHN418** | 27/60 | 45 |
| **15B/SH8286** | 15/54 | 28 |
| **23F/P1121** | 2/16 | 13 |
| **23F/P833** | 3/60 | 5 |

<sup>a</sup> Colonization was defined as growth of the specified pneumococcal strain at any time point on Columbia blood agar from nasal wash.
<sup>b</sup> Colonization efficiency was defined as the percentage of challenged volunteers who were positive for the challenge strain at any time point, independent of the dose received.

### Experimental colonization density and duration upon human challenge are comparable between pneumococcal strains

Colonization densities (Figure 2) and duration of colonization (Table 3) of 6B/BHN418, 15B/SH8286, 23F/P833 and 23F/P1121 were assessed over a two-week study period and compared using a general linear mixed model. Densities and durations of colonization episodes of 23F/P1121 and 23F/P833 were excluded from the analysis due to the low number of colonized individuals (n=2 and n=3, respectively).

**Table 3.** Duration^a^ of experimental human pneumococcal colonization.

| **Pneumococcal strain** | **2 days** | **7 days** | **14 days** |
| --- | --- | --- | --- |
| **6B/BHN418** | 2/27 (7.4%) | 7/27 (25.9%) | 18/27 (66.6%) |
| **15B/SH8286** | 3/15 (20%) | 3/15 (20%) | 9/15 (60%) |
| **23F/P1121** | 0/2 (0%) | 0/2 (0%) | 2/2 (100%) |
| **23F/P833** | 1/3 (33.3%) | 0/3 (0%) | 2/3 (66.6%) |

<sup>a</sup> Duration defined as number of volunteers who were colonised for 2, 7 or 14 days.

**Figure 2:**
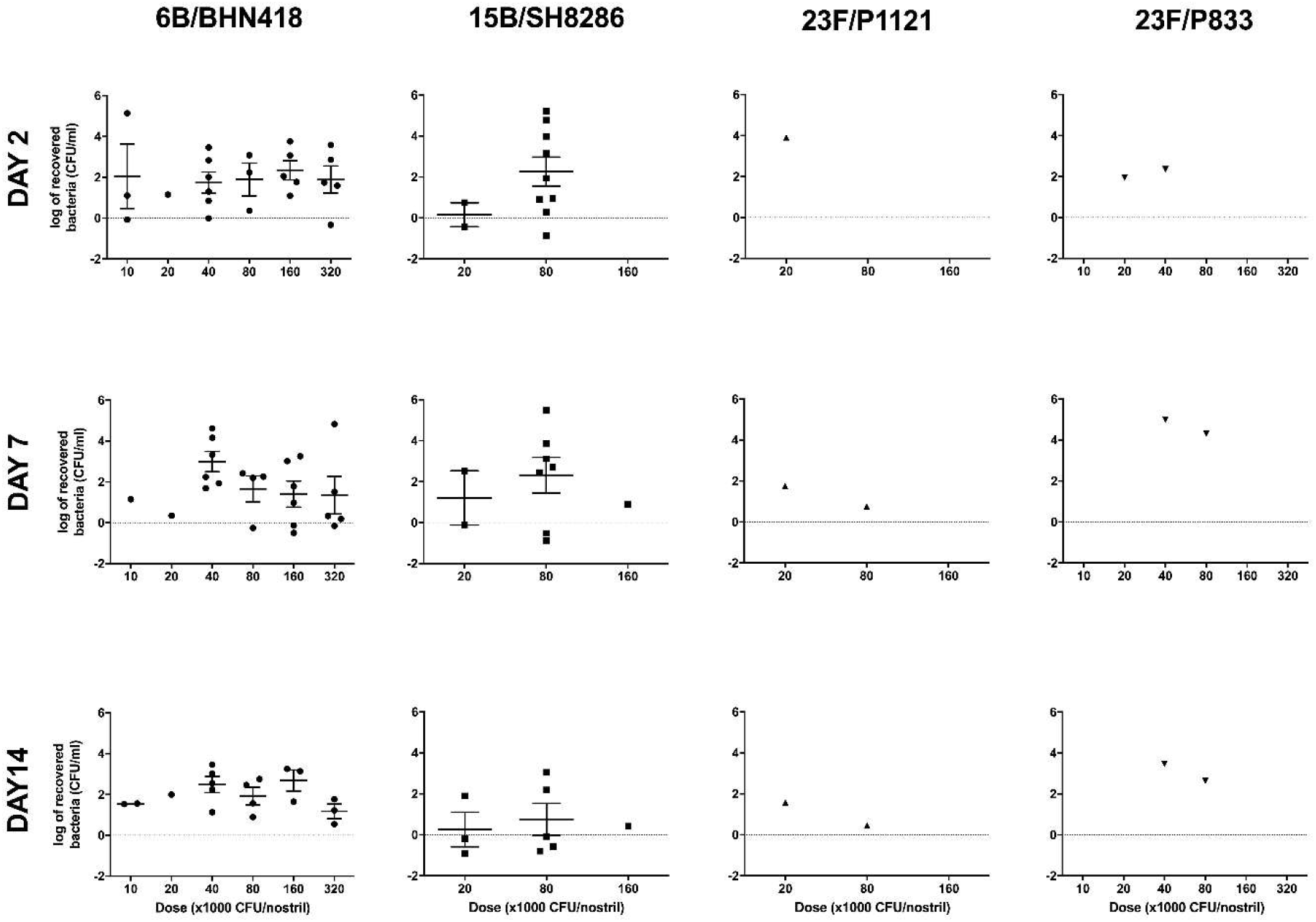
Pneumococcal colonization densities are comparable between strains. Nasal washes were taken at 2, 7, and 14 days post-challenge to determine pneumococcal density. Nasal wash was serially diluted and pneumococcal colonies were quantified on Columbia blood agar. Density of those colonised, is reported as the log of colony forming units per ml of nasal wash (CFU/ml) returned. Data bars represent the mean ± SEM.

Colonization densities were comparable between 15B/SH8286 and 6B/BHN418 after adjusting for day and dose (P=0.4924). Similar the duration of colonization defined as the last day at which colonization could be detected by classical microbiology was comparable between those challenged with 6B/BHN418 and 15B/SH8286 after adjusting for dose (P=0.7009).

### The predictive value of the murine model for experimental human pneumococcal colonization

Colonization rates and densities of 6B/BHN418, 15B/SH8286, 23F/P833 and 23F/P1121 were evaluated in murine models of pneumococcal colonization in order to assess whether colonization in mice is predictive of colonization efficiency in the human challenge model (Figure 3A and 3B). In mice, 23F/P833 failed to achieve successful colonization in Swiss Webster mice (0/5 = 0%) and colonized 3 out of 6 (50%) C57BL/6 mice with low colonization density (2.5 × 10^1^ ± 7.0 CFU). Strains 6B/BHN418 and 15B/SH8286 (C57BL/6) as well as 23F/P1121 (Swiss Webster) all achieved successful colonization (5/5 = 100% for 6B/BHN418, 7/7 = 100% for 15B/SH8286 and 4/4 = 100% for 23F/P1121), with similar colonization densities amongst the strains (6B/BHN418: 7.4 × 10^3^ ± 2.1×10^3^ CFU; 15B/SH8286: 1.3 × 10^4^ ± 1.8 × 10^3^ CFU, 23F/P1121: 1.7 × 10^4^ ± 1.4 × 10^4^ CFU). Our results indicate murine colonization was not a good predictor of human experimental colonization efficiency.

**Figure 3:**
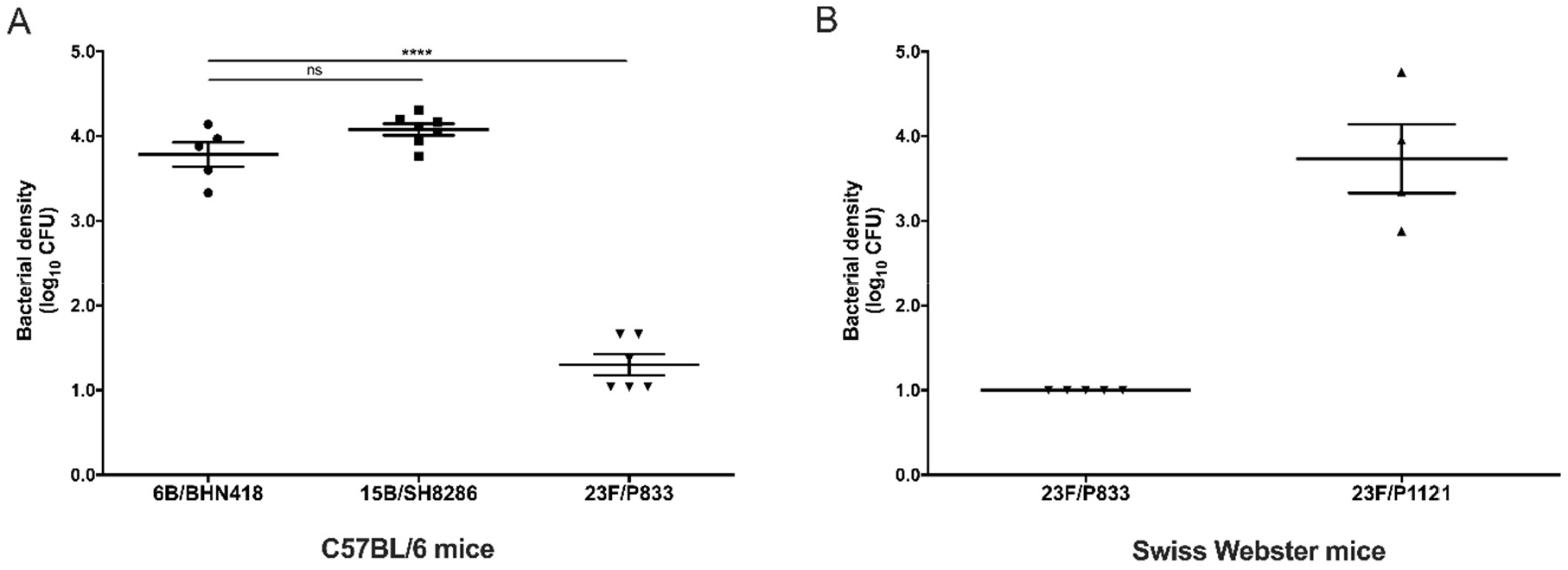
The predictive value of the murine model for experimental human pneumococcal colonization. (A) C57BL/6 mice were intranasally inoculated with 1 × 10^6^ CFU 6B/BHN418 (circle, n=5), 15B/SH8286 (square, n=7) or 23F/P833 (triangle apex down, n=6). Colonization densities were determined 5-days post-challenge. Data bars represent the mean ± SEM. P values using one-way ANOVA with Dunnett’s post hoc analysis *P ≤ 0.05, ** P ≤ 0.01, *** P ≤ 0.001, **** P ≤ 0.0001). (B) Swiss Webster mice were intranasally inoculated with 1 × 10^7^ CFU of 23F/P833 (triangle apex down, n=5) or 23F/P1121 (triangle apex up, n=4). Colonization densities were determined 5-days post-challenge. Data bars represent the mean ± SEM.

### Superior colonization efficiency of serotype 6B clone BHN418 upon human challenge is associated with pneumococcal chain length

Investigating the superior colonization efficiency of 6B/BHN418, we evaluated three key microbial properties of the pneumococcus. Strains 6B/BHN418, 15B/SH8286, 23F/P1121 and 23F/P833 were evaluated for non-opsonic neutrophil mediated killing (NMK, Figure 4A), the ability to adhere to primary human nasal cells (epithelial adherence, Figure 4B) and pneumococcal chain length (Figure 4C).

**Figure 4:**
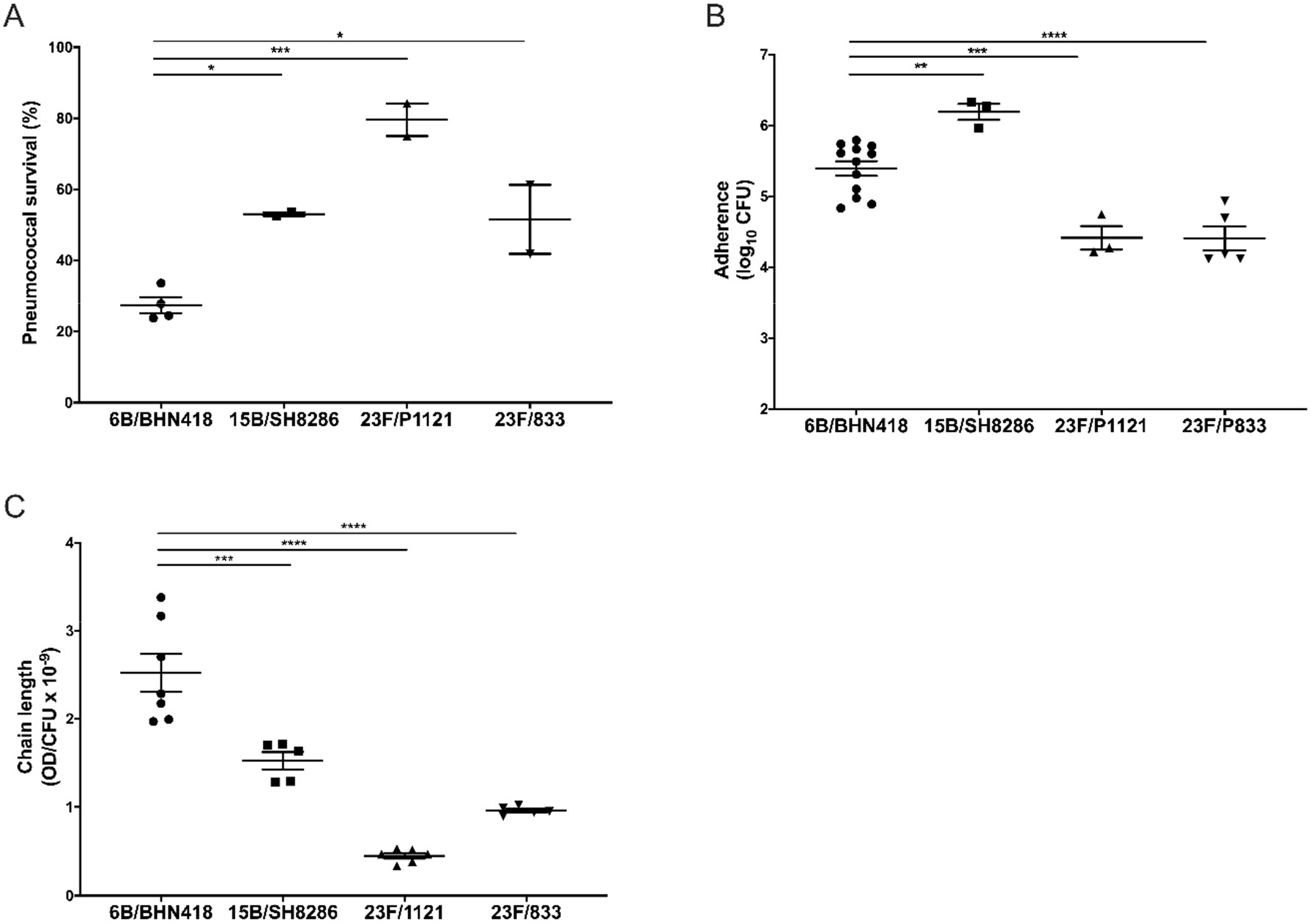
Superior colonization efficiency of serotype 6B clone BHN418 upon human challenge is associated with pneumococcal chain length. Pneumococcal strains 6B/BHN418 (circle), 15B/SH8286 (square), 23F/P1121 (triangle apex up) and 23F/P833 (triangle apex down) were evaluated for (A) non-opsonic neutrophil mediated killing (B) epithelial adherence and (C) average chain length. Bars represent the mean ± SEM. P values using one-way ANOVA with Dunnett’s post hoc analysis *P ≤ 0.05, ** P ≤ 0.01, *** P ≤ 0.001, **** P ≤ 0.0001).

Evaluation of NMK showed significant differences between the tested strains (ANOVA, P=0.0008). In contrast to the human challenge data, we observed the lowest level of NMK (27% ± 2.2) with the most efficient colonizer (6B/BHN418). Significantly higher levels of survival could be observed with 15B/SH8286 (53 ± 0.6 %, P=0.0140) and 23F/P833 (52 ± 9.7 %, P=0.0184). Pneumococcal strain 23F/P1121 showed the highest level of survival (80 ± 4.6 %) and was significantly more likely to survive (P=0.0004) when compared to 6B/BHN418. Evaluation of epithelial adherence showed significant differences between the isolates (ANOVA, P<0.0001). In contrast to the human challenge data, we observed the highest level of epithelial adherence (mean ± SEM) with 15B/SH8286 (1.7 × 10^6^ ± 3.7 ×10^5^ CFU), while 6B/BHN418 (3.2 × 10^5^ ± 5.8×10^4^ CFU) showed significantly (P=0.0052) lower levels of adherence. Epithelial adherence of 23F/P833 (3.6 × 10^4^ ± 1.5 × 10^4^ CFU) and 23F/P1121 (3.1 × 10^4^ ± 1.3 × 10^4^ CFU) were significantly lower when compared to 6B/BHN418 (P<0.0001 and P=0.0008, respectively).

Evaluation of average pneumococcal chain length showed significant differences between the tested strains (ANOVA, P<0.0001). The highest ratio of OD/CFU (mean ± SEM) and therefore larger chains were observed with 6B/BHN418 (2.53 ± 0.22 ×10^−9^). Significantly smaller chains were observed for 15B/SH8286 (1.53 ± 0.10 ×10^−9^, P=0.0002). Average chain length of 23F/P833 (0.96 ± 0.02 ×10^−9^) and 23F/P1121 (0.45 ± 0.03 ×10^−9^) were significantly smaller when compared to 6B/BHN418 (P<0.0001). Visual assessment using microscopy confirmed a noticeable difference in magnitude and quantity of chains between 6B/BHN418, 15B/SH8286 and 23F clones (Supplementary Figure 1).

In summary, our data suggest the hypothesis that pneumococcal chain length is associated with experimental colonization efficiency.

## Discussion

Using the EHPC model, we have shown that colonization rates varied greatly depending on the strain used for challenge. Serotype 6B clone BHN418 was the most efficient colonizer in this model. Other studies carried out using the EHPC model and involving challenge with this strain have shown stable overall rates of colonization (ES (95% Cl): 0.47 (0.43,0.52)) (Rylance et al., 2019, Jochems et al., 2017, Skoberne et al., 2016, Collins et al., 2015, Ferreira et al., 2013). Previous reports of pneumococcal colonization densities amongst different serotypes have been controversial (Baggett et al., 2017, Rodrigues et al., 2016). Moreover, comparison of pneumococcal duration during natural colonization events presents a challenge as the exact onset of colonization is difficult to determine. Here, we compare colonization densities and duration of different pneumococcal isolates upon human pneumococcal challenge over a 14- day period. Our data, show no significant differences of colonization density and duration between the tested pneumococcal strains.

Murine models have historically been used to study pneumococcal colonization behaviour. Here we show with four strains that colonization of mice is not predictive of pneumococcal colonization efficiency in the human model. Despite successfully establishing colonization and achieving similar colonization densities as serotype 6B clone BHN418 in mice, serotype 15B clone SH8286 as well as serotype 23F clone P1121 colonized the human host less efficiently. Among many potential reasons to explain the lack of concordance between murine and human models of colonization, two important ones stand out: first, mice are not pneumococcus’ natural host and secondly, unlike adults and children as they age, laboratory mice are predominantly immunologically naïve to the bacterium.

As new challenge models are setup further knowledge of microbial factors influencing colonization efficiency is of great relevance. The relevance of neutrophil driven activity against the pneumococcus during the early stages of colonization has been previously demonstrated (Jochems et al., 2017, Ronchetti et al., 2002, Nikolaou et al., 2018). Further, resistance to non-opsonic NMK has been shown to correlate with higher colonization prevalence, highlighting the potential role of neutrophils in driving colonization patterns (Weinberger et al., 2009). In contrast, our data does not support a positive link between resistance to non-opsonic NMK and colonization efficiency in the human model. In fact, we observed the lowest level of resistance with the most efficient colonizer. This negative association may highlight the different role of the pneumococcal capsule in phagocytosis and epithelial binding. Yet, *in vivo* NMK is often aided by opsonisation of the bacteria by antibodies against the capsule or other surface antigens which could affect acquisition of the pneumococcus and in turn colonization efficiency.

Pneumococcal adherence to the nasal epithelium has been described as the last step in establishing stable colonization (Kadioglu et al., 2008, Siegel and Weiser, 2015, Weiser et al., 2018). Recent data investigating the role of pneumococcal sensing at the human epithelium demonstrated microcolony formation at the mucosa as early as 2 days after pneumococcal challenge (Weight et al., 2019), highlighting the potential importance of this step during establishment of colonization. Animal models as well as *in vitro* models have further demonstrated that the ability of the bacterium to adhere depends on serotype (Nelson et al., 2007, Trzcinski et al., 2015, Weinberger et al., 2009). In contrast, our data does not support an association of *in vitro* epithelial adherence and the ability of the pneumococcus to colonize using the EHPC model.

The formation of pneumococcal chains has previously been indicated to provide a competitive advantage during colonization (Rodriguez et al., 2012, Young, 2006). Here, we evaluated average chain length of four pneumococcal strains used in the human challenge model and showed that superior colonization efficiency is associated with a high magnitude of average pneumococcal chain length. This agrees with work by Weiser and colleagues showing that increased chain length of pneumococcus promotes adherence and colonization in *in vitro* human cell lines and murine models (Rodriguez et al., 2012). Increased chain length is associated with great surface area per particle, which could promote adherence to host surfaces, although not observed here with primary nasal cells in culture. Based on our data, we propose the hypothesis that chain length is related to the ability of a pneumococcal strain to colonize in the human challenge model.

Here, we investigated the superior colonization efficiency of strain 6B/BHN418 by comparing three of its microbial properties with strains demonstrating lower efficiency upon human challenge. Of note, strain 23F/P1121 was obtained following experimental human colonization with strain 23F/P833 in an earlier study (McCool et al., 2002). Although these strains are otherwise isogenic, passage in the human nasopharynx resulted in the acquisition of genetic polymorphisms. Such genetic differences could account for the observed differences of serotype 23F clone P1121 from its parent strain, including decreased NMK, increased murine colonization and shorter chain length. The properties tested here are not exclusive and other properties influencing colonization efficiency may exist. Nevertheless, as new pneumococcal challenge models are setup we recommend to give preference to those isolates with chain length equal or greater than that of serotype 6B clone BHN418 (OD/CFU ≥ 2.53 ± 0.22 ×10^−9^).

The role of host immunity in the establishment of colonization has been highlighted by us and others. We have previously reported that high baseline levels of blood capsular specific IgG memory B-cells but not IgG levels associate with protection against acquisition of the serotype 6B clone BHN418 pneumococcus in the human challenge model (Pennington et al., 2016). Moreover, IgG levels to several pneumococcal proteins did not correlate with protection against experimental colonization (Ferreira et al., 2013). While our present study concentrated on the identification of microbial characteristics, it is unclear whether the lower efficiency of the other strains tested in the EHPC model may be associated with other host factors conferring immunity against these strains. We recently reported that epithelial sensing of the serotype 6B clone BHN418 is less pronounced and leads to lower inflammatory responses when compared to other strains *in vitro* (Weight et al., 2019).

Further important limitations of our study are the small number of pneumococcal strains tested and the follow up of volunteers for 14 days only. These limitations are mainly caused by the fact that human challenge models require substantial clinical and laboratory infrastructure to ensure volunteer safety as well as precise challenge and bacterial detection, which is costly and logistically demanding. This is the first study to compare multiple serotypes in experimental human pneumococcal colonization. Methods of stock preparation, volunteer selection and exposure have been standardised in our unit over a decade. The differences observed are therefore unlikely to be due to experimental factors but rather either pathogen or host determinants of colonization.

Here, we demonstrate differences in pneumococcal colonization efficiency upon human challenge and highlight the possibility that pneumococcal chain length is related to the ability of the bacterium to colonize experimentally in the human challenge model.

## Data Availability

Nucleotide sequence of the challenge isolate 6B/BHN418 can be accessed in GenBank under accession numbers ASHP00000000.1. Nucleotide sequence of the challenge strains 23F/P833, 23F/P1121, and 15B/SH8286 can be accessed in the European Nucleotide Archive (ENA) using the following accession numbers: for 23F/P833 - ERS743506, for 23F/P1121 - ERS1072059 and for 15B/SH8286 - ERS2632437. The source data for figures displayed in this paper are available from the corresponding author on request. This study did not generate new unique reagents. Further information and requests regarding resource availability concerning this study should be directed to the Lead Contact, Prof. Daniela Ferreira.

https://www.ncbi.nlm.nih.gov/nuccore/ASHP00000000.1

https://www.ebi.ac.uk/ena/data/search?query=ERS743506

https://www.ebi.ac.uk/ena/data/search?query=ERS1072059

https://www.ebi.ac.uk/ena/data/search?query=ERS2632437

## Acknowledgements

The authors thank Prof. Peter Hermans (Radboud University Medical Center) for donating the 6B/BHN418 pneumococcal strain, originally from Prof. Birgitta Henriques-Normark (Karolinska Institute). They also thank the safety committee: Prof. Rob Read (University of Sheffield), Prof. David Lalloo (Liverpool School of Tropical Medicine), and Dr. Brian Faragher (Liverpool School of Tropical Medicine) and the clinical team at the Royal Liverpool and Broadgreen University Hospitals NHS. The authors also thank all the volunteers for their participation. The work was supported by the Bill and Melinda Gates Foundation (GCE award to SBG, no: OPP1035281 and grant award to DMF OPP1117728), the Medical Research Council to SBG and DF (grant MR/M011569/1) and the National Institute for Health Research Comprehensive Local Research Network. The funders had no role in study design, data collection and analysis, decision to publish, or preparation of the manuscript. The study was co-sponsored by the Royal Liverpool and Broadgreen University Hospitals NHS trust and the Liverpool School of Tropical Medicine. JNW was supported by grants from the U.S. Public Health Service (RO1 AI05168 and RO1 AI38446). Confocal imaging facilities were funded by a Wellcome Trust Multi-User Equipment Grant (104936/Z/14/Z). Results were partially presented at EuroPneumo 2013 and 2019 and ISPPD 9 (2014).

## Methods

### Recruitment and ethical statements

Healthy adult non-smoking volunteers who had no close contact with at risk individuals (including young children and the elderly) were enrolled to an experimental human pneumococcal challenge (EHPC) trial. Ethical approval was obtained from the National Health Service Research Ethics Committee (11/NW/0592, 15/NW/0931). The inclusion age slightly varied between the studies with 18-60 for 11/NW/0592 and 18-50 for 15/NW/0931. All experiments conformed to the relevant regulatory standards (Human Tissue Act, 2004). Informed consent was obtained from all volunteers.

All animal experiments were performed in accordance with local and national ethical approval guidelines. The experimental protocol “RU-DEC2014-215” and “RU-DEC2015-0139” involving the challenge of C57BL/6 mice were approved by the Animal Ethics Committee of the Radboud University, Nijmegen, the Netherlands. All procedures involving Swiss Webster mice were reviewed and approved by the Institutional Animal Care and Use Committee at Tufts University School of Medicine, Boston, USA.

### Bacterial strains

Four different clinical strains comprising three pneumococcal serotypes were used in these experiments (serotype/clonal name): 6B/BHN418 (gifted by Prof. Peter Hermans, Radboud University Medical Center), 23F/P833 and 23F/P1121 (gifted by Prof. Jeffrey Weiser, NYU School of Medicine), and 15B/SH8286 (gifted by Dr David Cleary, University of Southampton) were used for challenge in humans. 23F/P1121 was originally isolated from the nasopharynx of subjects enrolled in a USA pneumococcal challenge trial (McCool et al., 2002).

### Stock preparation and challenge

Bacterial stock preparation and challenge were performed as previously described (Gritzfeld et al., 2013). Volunteers who received pneumococcus 6B/BHN418 or 23F/P833 were subdivided into six groups of 10. Each of the six groups received a different challenge dose, starting at 1×10^4^ CFU/naris and doubling up to 32×10^4^ CFU/naris. Volunteers who received 23F/P1121 and 15B/SH8286 were subdivided into three groups each receiving a challenge dose of 2×10^4^ CFU/naris, 8×10^4^ CFU/naris, and 16×10^4^ CFU/naris, respectively.

### Nasal washing and detection of pneumococcal density

Nasal wash samples were collected before challenge and at days 2, 7 and 14 post challenge and were processed as previously described (Gritzfeld et al., 2013). Briefly, Columbia blood agar plates (PB0122A, Oxoid/Thermo Scientific) were inspected following 24 hours incubation at 37°C, 5% CO_2_. Alpha haemolytic, draughtsman-like colonies were sub-cultured for optochin sensitivity and bile solubility. Latex agglutination was used to confirm pneumococcal serotype. If pneumococci were detected at any nasal wash time point, the volunteer was considered colonization positive.

Pneumococcal density was determined as previously described with minor modifications (Gritzfeld et al., 2013). Briefly, 100 μl of skim-milk tryptone glucose glycerol (STGG) medium was added to the nasal wash bacterial pellet and the total volume of pellet plus STGG was determined. The pellet plus STGG was then serially diluted and plated on Columbia blood agar. The next day CFU per μl were determined and multiplied by the volume of the pellet plus STGG. This value was then divided by the amount of nasal wash returned by the volunteer to get CFU/ml of nasal wash.

### Murine models of colonization

Bacterial cultures were grown in THY broth to mid-exponential phase and adjusted to the desired concentration with PBS. Intranasal challenge was performed in anesthetized (isoflurane) 7-week old C57BL/6 mice (Charles River Laboratories, Germany) (5 μl/naris; 10^6^ CFU) or in anesthetized (isoflurane) 6 to 9 weeks old Swiss Webster mice (Taconic Laboratories, USA) (5 μl/naris; 10^7^ CFU). Five days post-challenge, mice were euthanized, and the nasal cavity was flushed with 500 μl PBS per naris. Lavage was collected and plated for quantitative culture. The lower limit for detection was 11 CFUs for C57BL/6 mice and 10 CFUs for Swiss Webster mice.

### Pneumococcal adherence assay

Primary nasal cells were collected and processed as previously described (Jochems et al., 2017). Briefly, cells were dislodged from a curette by repeated pipetting and spun down at 440 × g for 5 min. Afterwards cells were cultured for a maximum of 27 days as described elsewhere (Muller et al., 2013). For the assay cells were seeded in duplicates at a minimum of 1×10^5^ cells per well and grown for 4-7 days. On the day of the assay cells were washed 3 times with HBSS^+/+^. Bacteria were prepared at 5×10^6^ CFU/ml in EMEM media (M5650, Sigma-Aldrich Co Ltd, Irvine, UK) supplemented with 1% heat-inactivated FBS (Fisher Scientific, Loughborough, UK) and 2mM L-Glutamin (Sigma-Aldrich Co Ltd, Irvine, UK). 1×10^6^ CFU of bacteria were added to the cells, centrifuged at 200g for 1min and placed into a tissue cell incubator (37 °C, 5% CO2, >90% relative humidity) for 3h. After 3h the bacterial containing liquid was removed and cells were washed 3 times with ice-cold HBSS^+/+^. To dislodge cells from the culture plate 0.25% Trypsin (Fisher Scientific, Loughborough, UK) was added for 10min at 37 °C. Cells were collected in an Eppendorf tube before washing the culture plate thoroughly and collecting any additional liquid. To permeabilise cells 1% Saponin was added to the collected liquid for 10min at RT. Thereafter liquid was vortexed and bacteria were quantified by plating serial dilutions onto Columbia blood agar plates (PB0122A, Oxoid/Thermo Scientific) followed by incubation in a tissue incubator (37 °C, 5% CO2, >90% relative humidity) over night. Results were expressed as colony forming units (CFU) and corrected for dose. Adherence is defined as adherent and internalized pneumococci.

### Quantitative assessment of chain formation

Bacteria were grown as described elsewhere (Gritzfeld et al., 2013) using Vegitone (Sigma-Aldrich Co Ltd, Irvine, UK) as growth media. Optical density (OD_600nm_) was assessed using FLUOstar Omega microplate reader and Omega software (V 5.1 R2). Once bacteria reached mid-log phase they were plated onto Columbia blood agar and incubated in a tissue incubator (37 °C, 5% CO2, >90% relative humidity) over night. The degree of chain formation was expressed as ratio of optical density to colony forming units.

### Neutrophil surface killing assay

The neutrophil surface killing assay was adapted from a previous publication (Weinberger et al., 2009). Briefly, pneumococcus (grown to mid log-phase) were thawed, diluted to 5×10^3^ CFU/mL in saline, plated onto Columbia blood agar in 10 replicates and allowed to dry at RT. Neutrophils were isolated from peripheral blood as described elsewhere (Morton et al., 2016, Pennington et al., 2016) and diluted to 2×10^6^ cells/ml. Neutrophils were then overlaid, onto the bacterial spots and allowed to dry before incubating at 37° with 5% CO_2_. All Columbia blood agar plates were incubated in a tissue incubator (37 °C, 5% CO2, >90% relative humidity) over night and quantified the next day. Results were expressed as colony forming units (CFU) and corrected for dose.

### Statistical analysis

Graph and statistical analysis were performed using GraphPad prism version 5.0 (GraphPad software Inc, USA) and SAS/STAT^®^ Version 9.4 (SAS Institute Inc, USA). Data was log transformed where appropriate. Normal distribution was analysed using Kolmogorov-Smirnov test. For *in vitro* data outliers were identified using R (V3.6.2) and removed from the data set. Significance levels were determined using general linear mixed model, chi-quare test or one-way ANOVA with Dunnett’s post-hoc analysis. All P values are two-tailed and considered significant if P ≤ 0.05. Data is represented as mean ± SEM.

### Resource availability

All clinical isolates used in humans were sequenced on a PacBio platform in a reference laboratory. Nucleotide sequence of the challenge isolate 6B/BHN418 can be accessed in GenBank under accession numbers ASHP00000000.1. Nucleotide sequence of the challenge strains 23F/P833, 23F/P1121, and 15B/SH8286 can be accessed in the European Nucleotide Archive (ENA) using the following accession numbers: for 23F/P833 - ERS743506, for 23F/P1121 - ERS1072059 and for 15B/SH8286 - ERS2632437. The source data for figures displayed in this paper are available from the corresponding author on request. This study did not generate new unique reagents. Further information and requests regarding resource availability concerning this study should be directed to the Lead Contact, Prof. Daniela Ferreira.

